# Homologous protein domains in SARS-CoV-2 and measles, mumps and rubella viruses: preliminary evidence that MMR vaccine might provide protection against COVID-19

**DOI:** 10.1101/2020.04.10.20053207

**Authors:** Adam Young, Bjoern Neumann, Rocio Fernandez Mendez, Amir Reyahi, Alexis Joannides, Yorgo Modis, Robin JM Franklin

## Abstract

The COVID-19 disease is one of worst pandemics to sweep the globe in recent times. It is noteworthy that the disease has its greatest impact on the elderly. Herein, we investigated the potential of childhood vaccination, specifically against measles, mumps and rubella (MMR), to identify if this could potentially confer acquired protection over SARS-CoV-2. We identified sequence homology between the fusion proteins of SARS-CoV-2 and measles and mumps viruses. Moreover, we also identified a 29% amino acid sequence homology between the Macro (ADP-ribose-1’’-phosphatase) domains of SARS-CoV-2 and rubella virus. The rubella Macro domain has surface-exposed conserved residues and is present in the attenuated rubella virus in MMR. Hence, we hypothesize that MMR could protect against poor outcome in COVID-19 infection. As an initial test of this hypothesis, we identified that 1) age groups that most likely lack of MMR vaccine-induced immunity had the poorest outcome in COVID-19, and 2) COVID-19 disease burden correlates with rubella antibody titres, potentially induced by SARS-CoV2 homologous sequences. We therefore propose that vaccination of ‘at risk’ age groups with an MMR vaccination merits further consideration as a time appropriate and safe intervention.

## 1. Introduction

The race to discover a vaccine to slow the evolving wave of coronavirus disease 2019 (COVID-19) is a crucial aspect in establishing control in what is proposed to be one of the world’s worst pandemics [Ferguson et al., 2020]. As the rate of infection escalates and the number of deaths continue to rise, any intervention that will decrease the spread of the virus, provide immunity or reduce the severity of the disease in people that developed potentially life-threatening symptoms cannot come early enough.

The striking feature of COVID-19 is that it has a disproportionate impact on the elderly population, yet the paediatric populations in many reports escape with mild, if any, symptoms [Ferguson et al., 2020]. This is peculiar in a viral illness, as one would assume that the collective immunity accumulated over a lifetime would arm elderly populations with better defences than their progeny. A study by the Centers for Disease Control and Prevention (CDC) found that children are most likely and people 65 and older least likely to get sick from influenza. Median incidence values (or attack rate) by age group were 9.3% for children 0-17 years, 8.8% for adults 18-64 years, and 3.9% for adults 65 years and older [Tokars et al., 2018]. A simple answer to why COVID-19 is different could be that the elderly population do not have the physiological reserve or regeneration capacity to fight such a severe burden of disease. However, more intriguingly, the vaccination history of the individual patient might contribute to the severity of the disease.

The triple vaccine containing attenuated measles, mumps and rubella (MMR) viruses was introduced in the early 1970s [Ministry of Health, NZ, 2018]. This was driven by large pandemics of the viruses for which individual vaccinations were made available in the late 1960s [Hamborsky et al., 2015]. In the case of rubella, between 1962 and 1965 a pandemic, originating in Europe, spread to the United States leading to an estimated 12.5 million cases which caused significant disability to the foetus of pregnant mothers resulting in blindness, deafness and death [Ministry of Health, NZ, 2018]. Currently, the MMR vaccination is given in two doses and is offered to children between 9 and 15 months initially with a second dose at 15 months to 6 years of age [World Health Organisation, 2017]. The vaccination suffers from reduced uptake as a result of falsified claims of a link to autism [Editors of The Lancet, 2010]. There are, however, wide differences between national health systems and their sub-regions in the years when they formally introduced each vaccine in their childhood vaccination schedules, or when they established national campaigns to promote specific vaccines for vulnerable population [Hamborsky et al., 2015; World Health Organisation, 2019].

Recent pre-print publications have suggested a link between Bacillus Calmette–Guérin (BCG) vaccination and possible acquired non-specific immunity against COVID-19 [Miller et al., 2020]. Whilst we accept that the BCG vaccination has the potential for providing non-specific immunity, we note that the vaccination statistics of each country against BCG would be similar to that of an MMR programme. Here we provide three preliminary lines of evidence to support the hypothesis that the MMR vaccine may provide protection against COVID-19.

## 2. Methods

### 2.1 Genomic and structural referencing paramyxovirus

Sequence homology searches (DELTA-BLAST search followed by iterated PSI-BLAST searches) of paramyxovirus sequences using SARS-CoV-2 as the query. A homology model of the postfusion structure of SARS-CoV-2 S2 based on the postfusion structure of SARS-CoV-1 S2 [Walls et al, 2017] was constructed to asses overall similarities in fold homology with a model of measles virus F based on the postfusion structure of human parainfluenza virus 3 (HPIV3) F [Yin et al, 2005].

### 2.2 Genomic and structural referencing of rubivirus

Iterated PSI-BLAST searches of rubivirus sequences with SARS-CoV-2 Orf1ab as the query were performed. The crystal structure of SARS-CoV-2 ADRP bound to ADP-ribose (Protein Data Bank ID 6W02) was used to generate a homology model of the rubella virus ADRP with the same overall folding.

### 2.3 Epidemiological data

Italy, Spain and Germany are the three European countries with the highest number of reported Covid-19 cases at the moment. Historic vaccination schedules or recommendations for these countries were identified from relevant bodies and the literature [Vaccination schedules in Spain, 2019; STIKO, Germany, 2020; Filia, 2003]. Average coverage for each 10-year age group was calculated using WHO / UNICEF coverage estimates for 1980-2018 [World Health Organisation, 2019]. Age-adjusted case-fatality risk was calculated, separately for males and females, as a percentage. This was done using data about the number of reported cases with confirmed Covid-19 for whom demographis were known, and the fatalities amongst them, which were obtained from national reports [State Regions Conference, Italy 2020; Istituto Superiore di Sanità 2020] and from the European Centre for Disease prevention and Control (ECDC) [European Centre for Disease, 2020].

### 2.4 Serology analysis

Serum samples were collected from anonymised patients admitted to Luton & Dunstable University Hospital under protocol REC: 20/YH/0125 approved by the NHS Health Research Authority. Cases were stratified into 2 groups based on severity. Moderate severity included patients admitted to hospital to receive level 1 care (ward based). The high severity category included patients requiring level 3 care (ITU/HDU). Rubella IgG levels were measured using VIDAS® RUB IgG (RBG) assay (Biomerieux, Lyon, France).

## 3. Results & Discussion

### 3.1 Homologous protein domains in SARS-CoV-2 and measles, mumps and rubella viruses

Given known similarities in the glycoprotein folds of paramyxoviruses and coronaviruses [Walls et al., 2017], we undertook amino acid sequence homology searches for measles and mumps viruses, as well as rubella, the third component of the MMR vaccine. Intriguingly, we identified homologous domains in severe acute respiratory syndrome coronavirus 2 (SARS-CoV-2), the COVID-19 causing virus, with rubella virus as well as the two paramyxoviruses.

The homologous sequences in SARS-CoV-2 and rubella, aligned in PSI-BLAST [Altschul et al., 1997] with an expected value of 10^−78^, encode Macro domains in SARS-CoV-2 non-structural protein 3 (NSP3), a papain-like protease, and in rubella virus p150, a protease/methyltransferase. Macro domains can bind ADP-ribose-1’’-phosphate, an NAD metabolite, and have ADP-ribose-1’’-phosphatase (ADRP) activity. The presence of Macro domains with ADRP activity in coronaviruses, paramyxoviruses and alphaviruses has been noted previously [Snijder et al., 2003; Saikatendu et al., 2005; Eriksson et al., 2008]. A crystal structure of the SARS-CoV-2 Macro domain bound to ADP-ribose (Protein Data Bank entry 6W02) allows the substrate binding pocket and catalytic residues to be identified, by analogy with other Macro domains.

The Macro domains of SARS-CoV-2 and rubella virus share 29% amino acid sequence identity, suggesting they have the same protein fold. We generated an atomic model of the rubella virus Macro domain by threading the rubella sequence onto the SARS-CoV-2 Macro structure, guided by secondary structure predictions, with PHYRE2 [Kelley et al., 2015] (100% confidence score; Fig. 1A). Residues conserved in the SARS-CoV-2 and rubella Macro domains include surface-exposed residues and residues required for ADP-ribose binding and ADRP enzymatic activity in other coronaviruses (Fig. 1). Notably, ADRP activity is required for a mouse coronavirus, MHV-A59 (mouse hepatitis virus A59), to cause acute hepatitis. Mutation of a single residue in the MHV-A59 Macro domain (N1348A) abrogates ADRP activity [Eriksson et al., 2008]. This residue is conserved in the Macro domains of SARS-CoV-2 and rubella. Together these observations suggest that ADRP activity could be important in COVID-19 and rubella pathogenesis.

**Figure 1:**
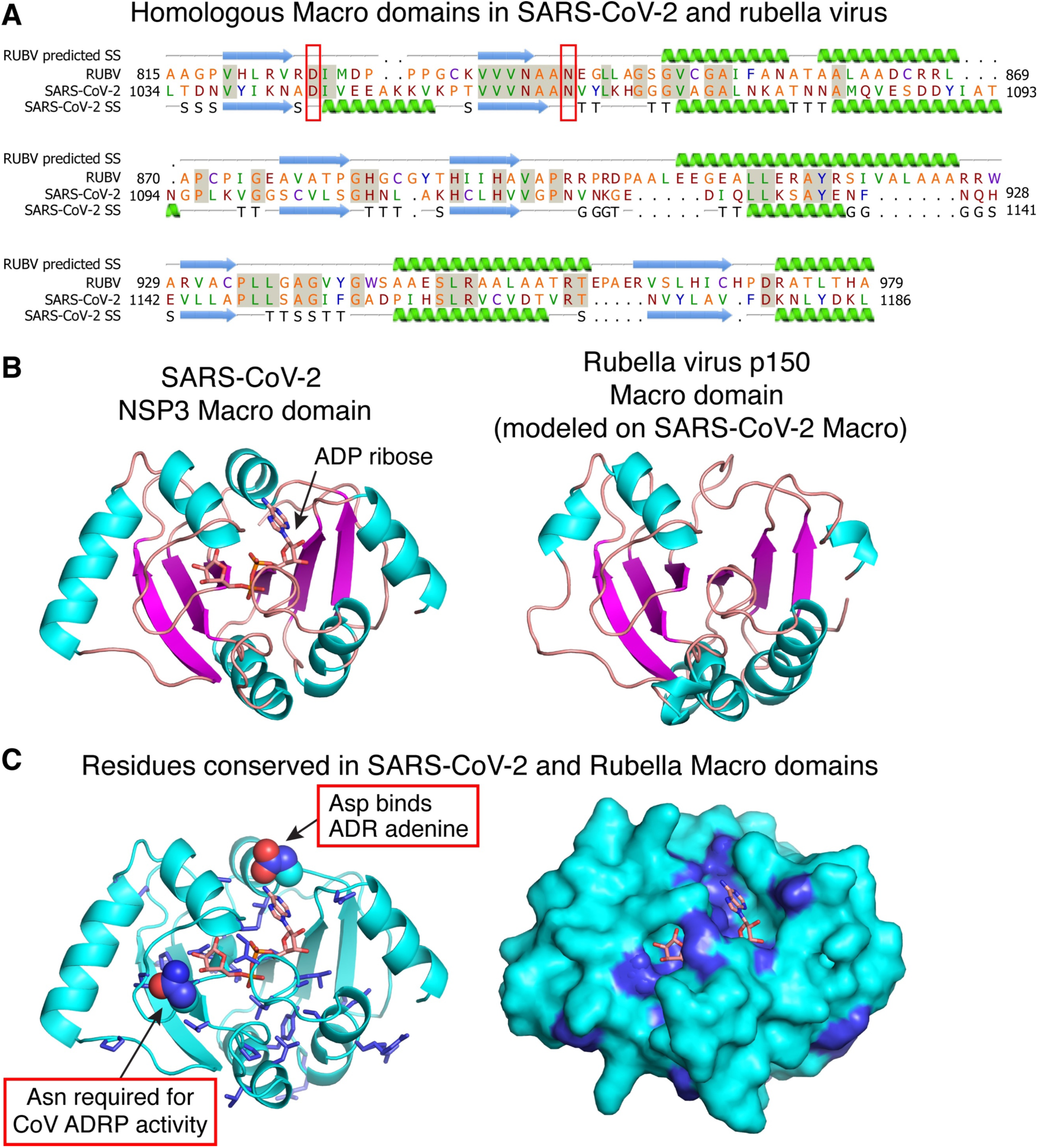
(**A**) Structure-guided sequence alignment and secondary structure prediction for the rubella virus Macro domain. The crystal structure of the SARS-CoV-2 Macro domain bound to ADP ribose was used as the template (Protein Data Bank entry 6W02). SS, secondary structure. (**B**) Left, structure of the SARS-CoV-2 Macro domain bound to ADP-ribose (ADR); Right homology model of the rubella virus Macro domain generated by one-to-one threading in PHYRE2^11^. (**C**) Amino acids conserved in SARS-CoV-2 and rubella Macro domains are shown in blue. Conserved residues include surface-exposed residues and residues involved in ADP-ribose phosphatase (ADRP) catalysis.

The Macro domain is present in the attenuated rubella virus used in the MMR vaccine. Although it is within a cytoplasmic non-structural protein, it could contribute to vaccine antigenicity if it released upon cell lysis, or displayed via MHC-I in vaccinated patients. Hence, we hypothesise that potentially immunogenic elements of the SARS-CoV-2 Macro domain that are conserved in rubella virus could serve as targets for B-or T-cell responses in patients who have had rubella or received a version of the vaccine. In this regard, we note certain antibodies raised against the S surface glycoprotein of SARS-CoV cross-neutralize SARS-CoV-2 [Wang et al., 2020; Lv et al., 2020; Walls et al., 2020], but it remains unclear to what extent non-neutralizing antibody cross-reactivity may lead to antibody-dependent enhancement [Lv et al., 2020].

Viral envelope glycoproteins catalyse an essential step in cell entry, fusion of the viral and cellular membrane. Coronavirus Spike glycoproteins are class I viral membrane fusion proteins. As such they share the same trimeric a-helical coiled-coil architecture as other class I fusogens including those from ortho- and paramyxoviruses [Walls et al., 2017; Skehel et al., 2000]. Structural homology searches with DALI [Holm et al., 2016] and molecular modelling with PHYRE2 [Kelley et al., 2015] confirmed clear structural homology between the SARS-CoV-2 Spike and paramyxovirus F proteins, including both measles and measles F in the postfusion conformation (DALI Z-score 6.3; Fig. 2). Homologous sequences in SARS-CoV-2 S2 and measles, aligned in PSI-BLAST [Altschul et al., 1997] with an expected value of 10^−101^, with 20% sequence identity over a 369-amino acid region (Fig. 3). An extensive set of surface-exposed residues is conserved in SARS-CoV-2 S2 and measles F.

**Figure 2:**
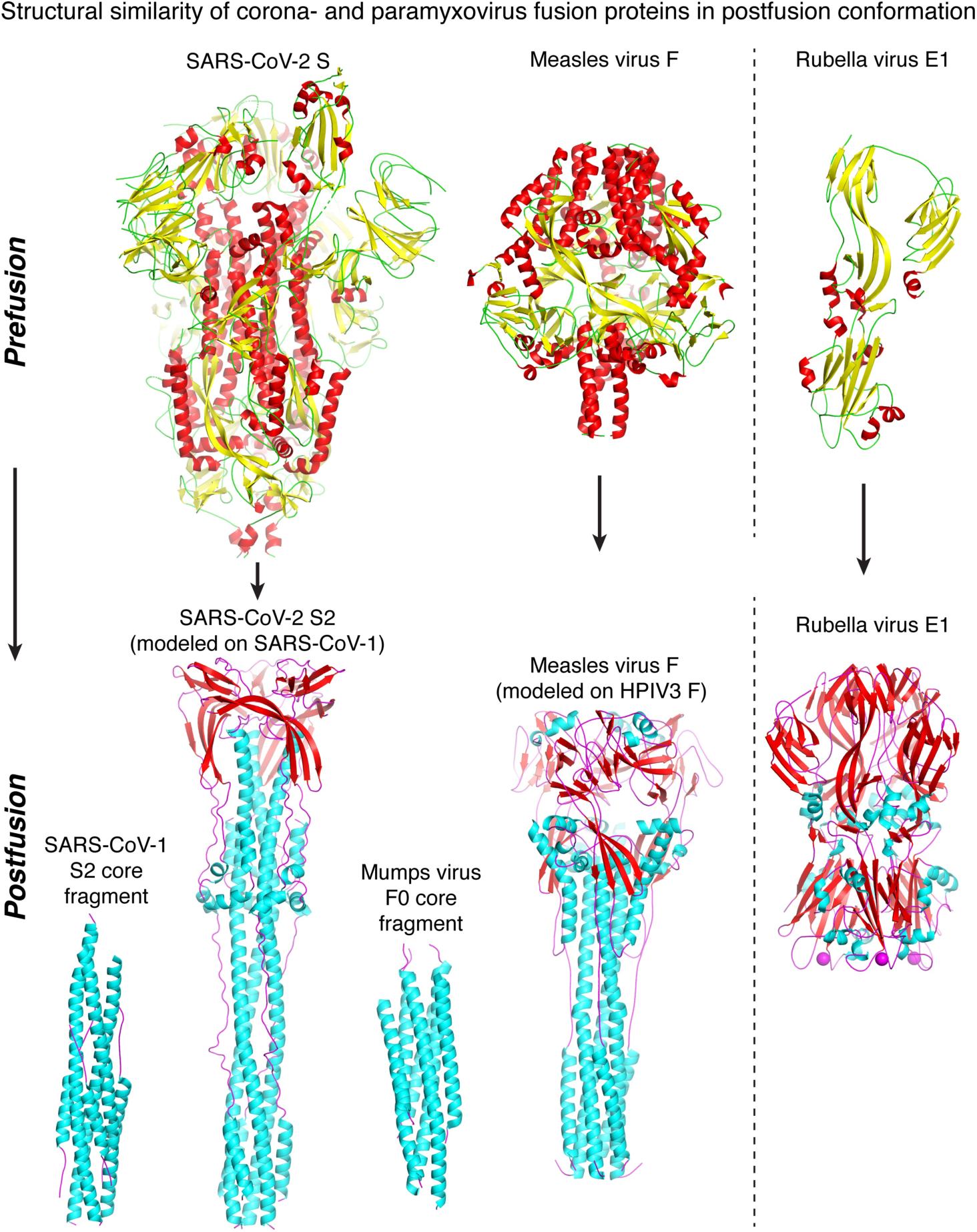
Coronavirus Spike (S) glycoproteins are class I membrane fusion proteins. SARS-CoV-2 S and paramyxovirus Fusion (F) proteins have different prefusion conformations (top left), but refold into similar ⍰-helical coiled-coil structures in their postfusion conformations (lower left). Rubella virus E1 is a class II fusion protein (right). Its pre- and postfusion folds are unrelated to those of class I fusion proteins. All proteins are drawn to scale. The following atomic models were used: prefusion SARS-CoV-2 S, PDB:6VSB; prefusion measles F, PDB:5YXW, prefusion rubella E1, PDB:5KHC; SARS-CoV-1 postfusion core, PDB:5ZVM, postfusion SARS-CoV-2, modelled with PHYRE2 based on PDB:6B3O (SARS-CoV1); postfusion mumps F0 core, PDB:2FYZ; postfusion measles F, modelled with PHYRE2 based on PDB:1ZTM; postfusion rubella E1, PDB:4ADI.

**Figure 3:**
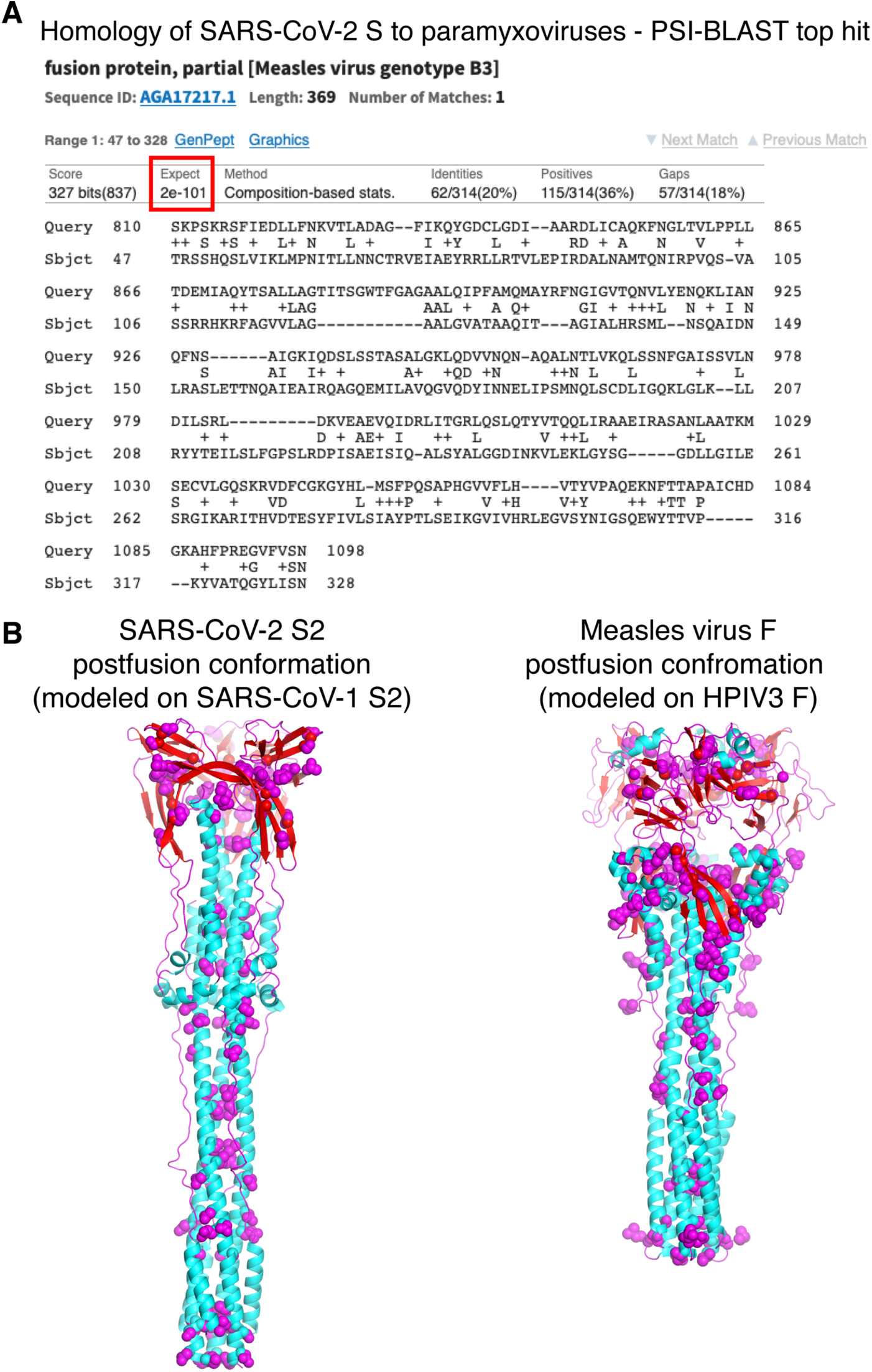
(**A**) Sequence homology searches (DELTA-BLAST search followed by iterated PSI-BLAST searches) of paramyxovirus sequences using SARS-CoV-2 as the query returned measles virus F as the top hit. (**B**) A homology model of the postfusion structure of SARS-CoV-2 S2 based on the postfusion structure of SARS-CoV-1 S2^15^ has a similar overall fold as a homology model of measles virus F based on the postfusion structure of human parainfluenza virus 3 (HPIV3) F^18^. Conserved in SARS-CoV-2 S2 and measles F based on the alignment in (A), shown in blue sphere representation.

### 3.2 MMR immunity and mortality risk in Covid-19

If the MMR vaccine provides protection against SARS-CoV-2, as the structural biology suggests, then one would predict a correlation between MMR vaccine coverage and age (and sex)-associated disease profiles of COVID-19. To test this prediction, we examined data from Italy, Spain and Germany, the three European countries with the highest number of reported Covid-19 cases at present. Historic vaccination schedules or recommendations for these countries were identified from relevant bodies and the literature [Vaccination schedules in Spain, 2019; STIKO, Germany, 2020; Filia, 2003; State Regions Conference, Italy 2020]. Average coverage for each 10-year age group was calculated using WHO / UNICEF coverage estimates for 1980-2018 [World Health Organisation, 2019]. Age-adjusted case-fatality risk was calculated, separately for males and females, as a percentage. This was done using data about the number of reported cases with confirmed Covid-19 for whom demographics were known, and the fatalities amongst them, which were obtained from national reports [Ministerio de Sanidad, Spain, 2020; Istituto Superiore di Sanità, 2020] and from the European Centre for Disease prevention and Control (ECDC) [European Centre for Disease, 2020].

Estimated average national immunisation coverages with a measles-containing vaccine in the studied countries ranged within 57-97% in Germany (1980-2018), 63-96% in Spain (1981-2017), and 53-89% in Italy (1990-2018). For a rubella-containing vaccine these ranges were the same as for a measles-containing vaccine in Italy and Spain, and for Germany it was 87-97% (1990-2018) [World Health Organisation, 2019] (Figure 1).

Germany, Spain and Italy began to introduce the rubella vaccine for pre-adolescent girls in 1972, 1972 and 1978, respectively [Vaccination schedules in Spain, 2019; STIKO, Germany, 2020; Filia, 2003]; and conducted vaccination campaigns for women in child bearing age from 1978, 1982 and the early 90s, respectively [Vaccination schedules in Spain, 2019; STIKO, Germany, 2020; Filia, 2003; State Regions Conference, Italy 2020]. Assuming the latter covered females up to 30 years old, this would cover women who are currently in the age group of 60-69 years old in Germany and Spain, and 50-59 years old in Italy. The introduction of MMR began in the 1976, 1981 and early 90s in Germany, Spain and Italy, respectively, for male and female infants [Vaccination schedules in Spain, 2019; STIKO, Germany, 2020; Filia, 2003; State Regions Conference, Italy 2020]. This covers males and females who are currently in the age group of 40-49 years old in Germany, 30-39 years old in Spain, and 20-29 years old in Italy.

Age-adjusted case-fatality risk related to Covid-19 disease increases with age in all countries. It has ranged within 0-10.1%, 0-21.5%, and 0-24% among females and 0-27.3%, 0-32.9%, and 0-30.8% among males, in Germany, Spain and Italy, respectively. In males, the age groups from which case-fatality risk started to be over 1% was 70-79, 50-59, 40-49 in Germany, Spain and Italy respectively. In females these age groups were 60-69 in Italy and Spain, and 70-79 in Germany (Fig. 4).

**Figure 4:**
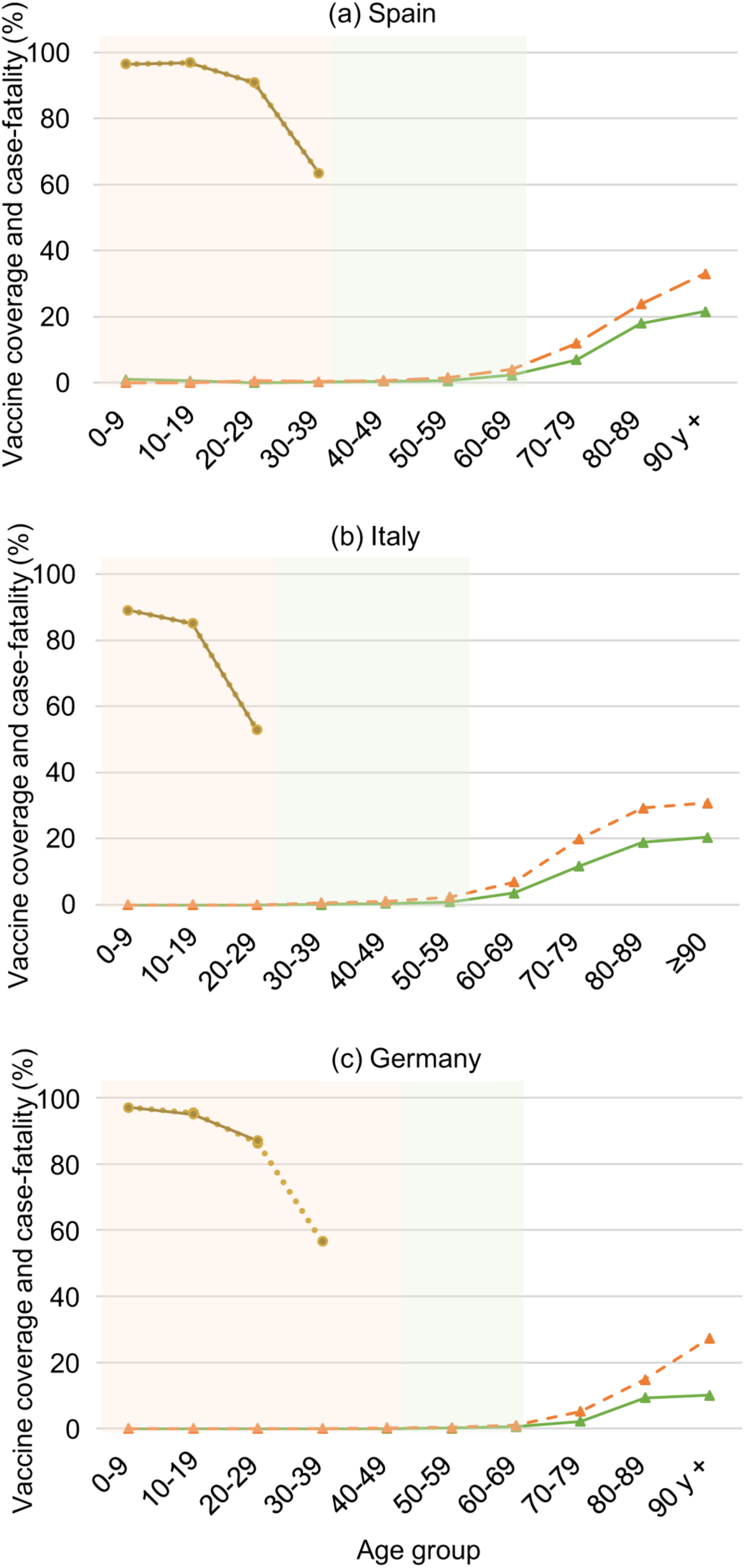
Measles and rubella vaccine coverages and Covid-19 age-specific case-fatality risk, by age group and sex, in (a) Spain, (b) Italy, and (c) Germany. Shaded background represents rubella vaccination target female population (green) and MMR vaccination target male and female population (orange), based on historical national vaccination schedules for compulsory and recommended vaccines and specific national vaccination campaigns.

We recognise that these data are, at this stage, preliminary and that there are a number of limitations, including the analysis being based on ecological rather than patient-level data, lack of reliable data on vaccine coverage and different ways of counting fatalities (people living in residencies not counted in some countries). Furthermore, there are differences in COVID-19 testing conditions, with mild suspect cases in Spain and Italy recommended to stay home and not tested, and no mass testing in any of the 3 countries studied. Nevertheless, older populations and males are both more likely to die from Covid-19, and less likely to be seropositive for rubella specific immunity, based on historical vaccination programmes of all three countries considered in this study. In order to conclude whether MMR vaccination can improve the outcomes from Covid-19 infection, a study using individual based data to compare MMR immunity status in the affected population is warranted.

### 3.3 Rubella IgG & IgM titres in COVID-19 patients correlate with disease burden

A further prediction of our hypothesis is that there should be a specific rise in rubella Immunoglobulin G (IgG) titres in COVID-19 patients, and that these should correlate with disease burden as a marker of immunogenicity against SARS-CoV2 [Kim et al., 2020]. To test this, serum samples were collected from patients admitted to Luton & Dunstable Hospital University Hospital (REC: 20/YH/0125) and stratified into 2 groups based on severity. Moderate severity included patients admitted to hospital to receive level 1 care (ward based). The high severity category included patients requiring level 3 care (ITU/HDU).

Patients with a high severity illness had on average increased levels of rubella IgG (161.9±147.6 IU/ml) compared to patients with a moderate severity of disease (74.5±57.7 IU/ml) (Fig. 5). In comparison, Immunoglobulin M (IgM) levels were 0.21±0.16 IU/ml in severe disease and 0.26±0.21 IU/ml in moderate disease. Whilst we accept that it is possible that this trend could be representative of pre-infection protection to rubella infection, it is not possible to determine this. In a study of 160 women of child bearing age, the IgG levels of non-infected patients measured between 24-143 IU/ml, suggesting that it is unlikely that those who developed severe symptoms of the disease had IgG levels far in excess of this prior to infection [Seker et al., 2004]. To test the specificity of this antibody response and to exclude the possibility that SARS-CoV2 infection was not driving a non-specific antibody response, we tested the six patients with the (3 from severe and 3 from moderate category) highest IgG titres and performed an ELISA against a varicella zoster virus epitope. All 6 patients returned positive samples ranging 1.86-3.46 (Ab Index) with reports in the literature of a response to a VZV vaccination being around 690 (Ab Index) [Smith-Norowitz et al., 2018]. This is consistent with the hypothesis that patients are specifically responding to SARS-CoV2 by identifying homology to rubella.

**Figure 5:**
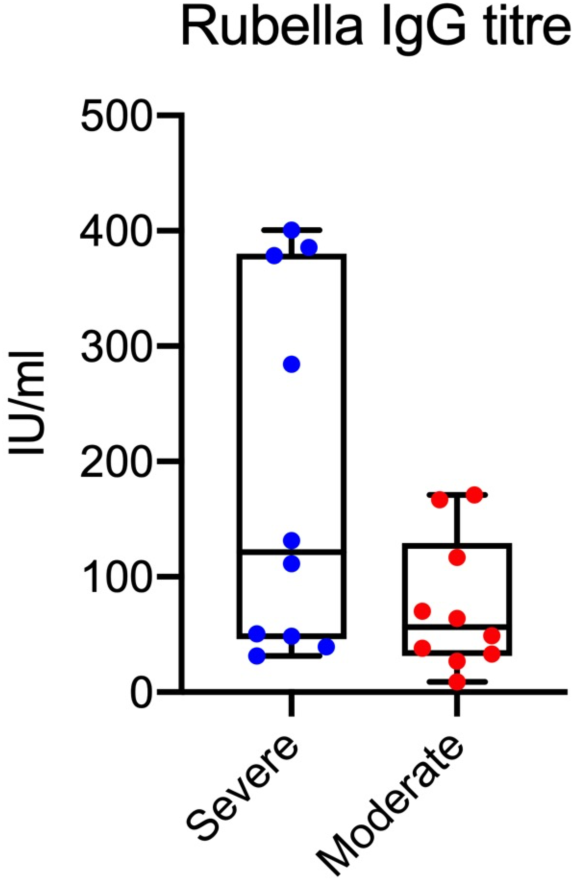
Rubella IgG titres (UI/ml) measured from COVID-19 positive patients. Patients were stratified into high and moderate severity groups based on the level of hospital care required.

The comparison of primary versus secondary infection responses in the literature describes similar elevation of IgG in secondary infection. Both rubella virus and dengue virus (DV) are pertinent examples of this. In primary infection, high levels of IgM are observed within a few days of disease onset, followed by a later production of IgG within a further few days at moderate levels [Alcon et al., 2002; Blacksell et al., 2008; Guzman & Kouri 1996; Koraka et al., 2001; Hamkar et al., 2005]. In secondary infection, IgM is detected a few days later and at lower levels than in primary infection, and IgG rapidly increases to very high levels [Guzman & Kouri 1996; Koraka et al., 2001; Hamkar et al., 2005]. This appears to resemble a similar pattern to our data where even in very high titres of IgG, IgM levels were barely raised.

In our case series, very high titres were discovered in patients who had been admitted for a period of less than 7 days. With this in mind, we suggest that IgG titters trend with disease burden on the basis of the shared homology between SARS-CoV2 and rubella virus. One proposal would be that the viral load, responsible for disease severity, is driving a high antibody response. Equally, however, as with DV, the presence of higher concentrations of antibodies could exacerbate disease burden if the antibodies were not neutralising in nature [Halstead, 2007]. Here the increased severity of secondary infections is believed to be a result, at least partially, from antibody-dependent enhancement (ADE) of DV infection, in which FcγR engagement by antibody-virus immune complexes facilitates virus entry into susceptible myeloid cell types [Halstead et al., 2003]. Interestingly, Balsitis et al., [2010] demonstrated that even when antibody levels were neutralising *in vitro*, cross-reactive polyclonal or monoclonal antibodies all enhanced disease *in vivo*. However, they also report complete elimination of viraemia with high-dose serotype-specific antibodies. Thus, we hypothesise that the administration of normal human immunoglobulin (NHIG) which contains antibodies against all three viruses would have the potential to rescue severe COVID-19 infection if administered in a timely fashion.

Taken together, our preliminary data would support the hypothesis that rubella vaccination could provide protection against poor outcome in COVID-19 infection. To determine if there is a potential effect of MMR vaccinations, it would be necessary to know the vaccination status of younger patients infected with SARS-CoV-2 and the severity of the disease. If there is a link, we propose that vaccination of ‘at risk’ age groups with an MMR vaccination should be considered as a time-appropriate and safe intervention. To create a SARS-CoV-2 vaccine will be arduous and may require time which we simply cannot afford. Meanwhile, some help could be immediately available to those in the greatest need.

## 4. Conclusion

SARS-CoV2 Spike glycoproteins are class I viral membrane fusion proteins that share structural similarities with the Fusion proteins from both measles and mumps viruses. The Macro domains of SARS-CoV-2 and rubella virus share 29% amino acid sequence identity. Interestingly, the residues conserved in the SARS-CoV-2 and rubella Macro domains include surface-exposed residues and are present in the attenuated rubella virus used in the MMR vaccine. We identified at a population level that both older populations and males are both more likely to die from COVID-19, and less likely to be seropositive for rubella-specific immunity, based on historical vaccination programmes of all three countries considered in this report. Finally, the hypothesis that this macro domain could be recognised by antibodies raised against rubella was supported by data that demonstrated that patients who have SARS-CoV2 infection had raised levels of rubella IgG to a level in keeping with secondary rubella infection. Taken together, we suggest that MMR will not prevent COVID-19 infection but could potentially reduce poor outcome. To conclude whether MMR vaccination can improve the outcomes from Covid-19 infection, a study using individual based data to compare MMR immunity status in the affected population is warranted.

## Data Availability

All data is is available

## Acknowledgements

The authors acknowledge the following support: Wellcome Trust Clinician PhD Fellowship to AY; Wellcome Trust Senior Research Fellowship 217191/Z/19/Z to Y.M.; The Adelson Medical Research Foundation (RF); the National Institute for Health Research (NIHR) Brain Injury MedTech Co-operative based at the University of Cambridge (RF, AJ - The views are those of the authors and not necessarily those of the NHS, the NIHR, or the Department of Health), and a core support grant from the Wellcome and MRC to the Wellcome-Medical Research Council Cambridge Stem Cell Institute (203151/Z/16/Z).

## Disclaimer

The views and opinions of the authors expressed herein do not necessarily state or reflect those of ECDC. The accuracy of the authors’ statistical analysis and the findings they report are not the responsibility of ECDC. ECDC is not responsible for conclusions or opinions drawn from the data provided. ECDC is not responsible for the correctness of the data and for data management, data merging and data collation after provision of the data. ECDC shall not be held liable for improper or incorrect use of the data.

